# Characterization of isolated human astrocytes from aging brain

**DOI:** 10.1101/2025.02.04.25321542

**Authors:** Geidy E. Serrano, Sidra Aslam, Jessica E. Walker, Ignazio S. Piras, Matthew J. Huentelman, Richard A. Arce, Michael J. Glass, Anthony J. Intorcia, Katsuko E. Suszczewicz, Claryssa I. Borja, Madison P. Cline, Sanaria H Qiji, Ileana Lorenzini, Suet Theng Beh, Monica Mariner, Addison Krupp, Rylee McHattie, Anissa Shull, Zekiel R Wermager, Thomas G. Beach

## Abstract

Astrocytes have multiple crucial roles, including maintaining brain homeostasis and synaptic function, performing phagocytic clearance and responding to injury and repair. It has been suggested that astrocyte performance is progressively impaired with aging, leading to imbalances in the brain’s internal milieu that eventually impact neuronal function and leads to neurodegeneration. Until now most of the evidence of astrocytic dysfunction in aging has come from experiments done with whole tissue homogenates, astrocytes collected by laser capture or cell cultures derived from animal models or cell lines. In this study we used postmortem-derived whole cells sorted with anti-GFAP antibodies to compare the unbiased, whole-transcriptomes of human astrocytes from control, older non-impaired individuals and subjects with different neurodegenerative diseases such as Parkinson’s disease (PD), Alzheimer’s disease (ADD) and progressive supranuclear palsy (PSP). We found hundreds of dysregulated genes between disease and control astrocytes. In addition, we identified numerous genes shared between these common neurodegenerative disorders that are similarly dysregulated; in particular, UBC a gene for ubiquitin, which is a protein integral to cellular homeostasis and critically important in regulating function and outcomes of proteins under cellular stress, was upregulated in PSP, PD, and ADD when compared to control.

## INTRODUCTION

Understanding the molecular events behind the normal physiological functions of astrocytes is crucial to unmask the roles that astrocytes may play in neurological disorders. The functions of astrocytes in the healthy adult brain are considered to involve potassium buffering, interstitial volume control, and maintenance of a low interstitial glutamate concentration. These functions are crucial for brain homeostasis and synaptic function [1–5]. Reactive gliosis, a component of neuroinflammation that involves structural and metabolic changes in astrocytes and microglia, is often a prominent feature of neurological disorders like Parkinson’s disease (PD), progressive supranuclear palsy (PSP) and Alzheimer’s disease dementia (ADD). Therefore, balanced astrocytic phagocytic clearance and response to injury and repair is crucial for the neuronal recovery following detrimental events. It has been suggested that astrocyte performance is progressively impaired with aging, leading to imbalances in the brain internal environment that eventually impact neuronal function and lead to neurodegeneration [1, 4, 6–9].

Nevertheless, until now the vast majority of evidence related to astrocytic dysfunction in aging and neurodegenerative disorders has come from experiments done with whole tissue homogenates, astrocytes collected by laser capture or cell cultures derived from animal models or cell lines [8, 10–13]. In this study we used an enriched population of whole human astrocytes to conduct an unbiased comparison of their gene expression in common neurodegenerative diseases and clinically normal subjects.

## METHOD DETAILS

### Subject selection and characterization

Fresh brain samples came from subjects who were volunteers in the Arizona Study of Aging and Neurodegenerative Disorders (AZSAND) and the Brain and Body Donation Program (BBDP; www.brainandbodydonationprogram.org), a longitudinal clinicopathological study of healthy aging, cognition, and movement in the elderly since 1997 in Sun City, Arizona [14, 15]. Cases for this study were selected based on their clinicopathological diagnosis, favoring those with the shortest postmortem intervals (PMI). Control subjects were cognitively unimpaired without parkinsonism or dementia and with lower amounts of AD pathology (CN; n=3). We also included a small number of cases without PD parkinsonism or cognitive impairment but with Lewy bodies in their brain, termed incidental Lewy body disease (ILBD; n=3). Impaired subjects included cases with PSP (n=5), ADD (n=4), and PD (n=6). Complete neuropathological examination was performed using standard AZSAND methods [15, 16]. Neurofibrillary degeneration was staged on the thick frozen sections by the original method of Braak [17, 18], and neuropathological ADD diagnoses were made when neuritic plaque densities and Braak stage met “intermediate” or “high” criteria according to NIA-AA criteria [19–22]. Non-ADD conditions were diagnosed using standard clinicopathological criteria, with international consensus criteria for those disorders where these were available [23, 24].

### Whole-cell-dissociated-suspension preparation

Whole-cell-dissociated-suspensions (WCDS) were generated from the prefrontal cortex of fresh human brain tissue as previously reported [25]. Briefly, fresh bilateral coronal sections of the frontal lobe were collected just anterior to the genu of the corpus callosum at autopsy. The grey matter was dissected and finely minced and incubated in Accutase (Innovative cell technologies BE cat#AT104) for 4 hrs all at 4°C, followed by mechanical disruption by repetitive pipetting. Homogenates were centrifuged and Accutase was replaced by Hank’s balanced salt solution (HBSS), following cell filtration using 100 and 70 µm filters. Myelin, neuropil, and other cellular debris were removed using 30% and 70% Percoll (GE Healthcare cat#17-0891-01). WCDS were stored in cryoprotectant solution (90%FBA and 10% DMSO + 1U/µl RNAse inhibitor) until sorting. Quality assessment for each suspension includes fixation and paraffin embedding of cell pellets that are subsequently cut at 3 um using a rotary microtome and stained with H&E and cell markers such as NeuN (Abcam cat#ab177487), MAP2 (Abcam cat#ab183830), and neurofilament (Abcam cat#ab8135); astrocyte marker GFAP(Millipore cat#MAB360 and Dako cat#Z0334; Figure 1); and microglia markers IBA1 (Wako cat#019-19741) and LN3 (Abcam cat#ab166777).

**Figure 1.**
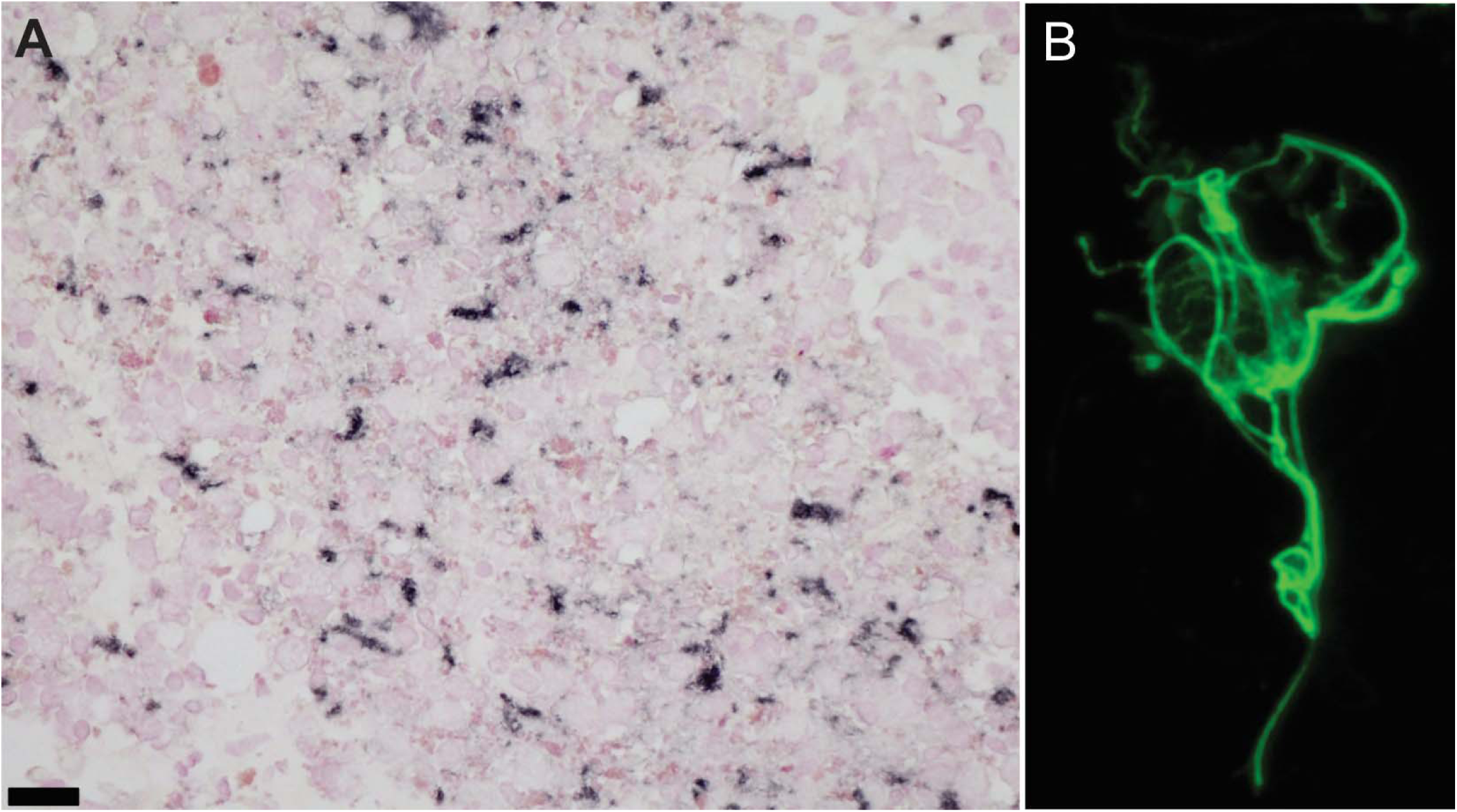
GFAP immunostains of astrocyte-enriched whole-cell-dissociated-suspensions pellets (A) showed an increase in the number of whole astrocytes (B) compared to other cell types.

### Astrocyte suspension enrichment

Frozen cryoprotected single-cell suspensions were rapidly thawed and fixed with cold 70% Methanol with 2mM EDTA for 30 minutes. Cells were magnetically sorted for GFAP using Dynabeads M-450 Epoxy (Thermo Fisher cat#14011) and GFAP 488 antibody (Abcam cat#194324). RNA was isolated from the sorted cells using Qiagen Rneasy micro kit (Qiagen cat#74004) by adding 100ul of RLT Lysis Buffer with B-Mercapoethanol to the bead-bound cells and then sonicating with light pulses (15 amps), placed in a magnetic rack for 1 minute and removing the supernatant into a new microcentrifuge tube and proceeding with RNA Isolation following manufacturer’s instructions. RNA yield and RNA Integrity (RIN) were calculated on RNA 6000 picochip (Agilent cat#5067-1513) using Agilent 2100 Bioanalyzer. The library was sequenced by 2X150 bp paired-end sequencing on an Illumina NovaSeq S4.

### Differential expression analysis

Sequencing reads were aligned to the Human Reference Genome GRCh38 using STAR [26] and summarized at the gene level using the HTSeq tool [27]. Quality controls were conducted using MultiQC [28] software. Outlier and batch effect detection were conducted through Principal Component Analysis (PCA), using R software v3.3.1. Samples with a total number of reads < 40M, and uniquely mapped reads < 55% were excluded from the downstream analysis. Genes with a total count of less than 10 were excluded. Normalization was performed using DEseq2. Gene expression differential analyses for every possible comparison between PSP, PD, ADD, ILBD, and control cases were performed using the R package DESeq2 [29], including age, gender, and PMI as covariates. The p-values were adjusted using the False Discovery Rate (FDR) method. Cell-specific gene set enrichment analysis was done using a list of 5,641 cell-specific genes generated from a brain single-nucleus RNA sequencing dataset from the DLPFC [30], as described in Piras et al. (2021)[31]. Genes from all cases were pulled together and the analysis was conducted using the R function enrichment (R package fgsea) adjusting for multiple testing with FDR. In addition, to gain insight into the biological functions of the DEGs, we performed a gene set enrichment analysis (GSEA) using the cluster Profiler package in R software [32].

### Hub gene identification

In order to identify astrocyte-related Hub genes commonly affected in PSP, PD and ADD, we used a search tool for the Retrieval of Interacting Genes (STRING) [33] to predict protein–protein interactions (PPI) of selected genes that were commonly dysregulated in all diseased cases when compared to CN. Cytoscape [34] was used to generate a network model from all genes with a confidence score ≥ 0.4 in STRING, by incorporating both physical and functional protein-protein interactions while CytoHubba, was used to identify Hub genes based on the connectivity of degree, DMNC, MCC, MNC, and closeness in the PPI network.

## Results

Cases with less than 40 million reads or with more than 55% of reads that could not be mapped to either the human or animal genome were eliminated. WCDS were enriched for astrocytes; this was confirmed visually with GFAP immunostains of the cell suspension pellets after sorting and by enrichment plots of *fGSEA* produced using *plotEnrichmnet* function (Figure 1).

There were no significant differences in group age means (p = 0.92). However, the youngest group (PD) had a mean age of 82, while for the oldest group (CN), the mean age was 89 (Table 1). PMI and RIN were also similar between groups (p = 0.17), varying from 2.9 to 4.4 hours and from 7.7 to 10.0, respectively. As expected, pathologies typical of ADD, PSP and LB pathology were significantly different between groups, with ADD having the highest densities of amyloid plaques and neurofibrillary tangles, PSP the highest level of pathological tau (summation of neuronal and glia tau scores) and PD with the highest densities of alpha-synuclein pathology (Table 1). Frontal cortex tissue adjacent to the sample collected for whole-cell suspensions stained for GFAP showed an increased number of reactive astrocytes in PSP cases, especially when compared to controls (Figure 2). Differential expression analysis was used to compare gene expression from each diseased group to CN. We found hundreds of dysregulated genes in each diseased group. PSP cases had the largest number of dysregulated genes when compared to CN (1,968), while ILBD only had one downregulated gene when compared to the CN group that has not yet been characterized by other studies (Table 2). A complete list of DEGs is reported in Supplement Table 1 and all data will be posted in Synapse (Project ID: syn64618348).

**Figure 2.**
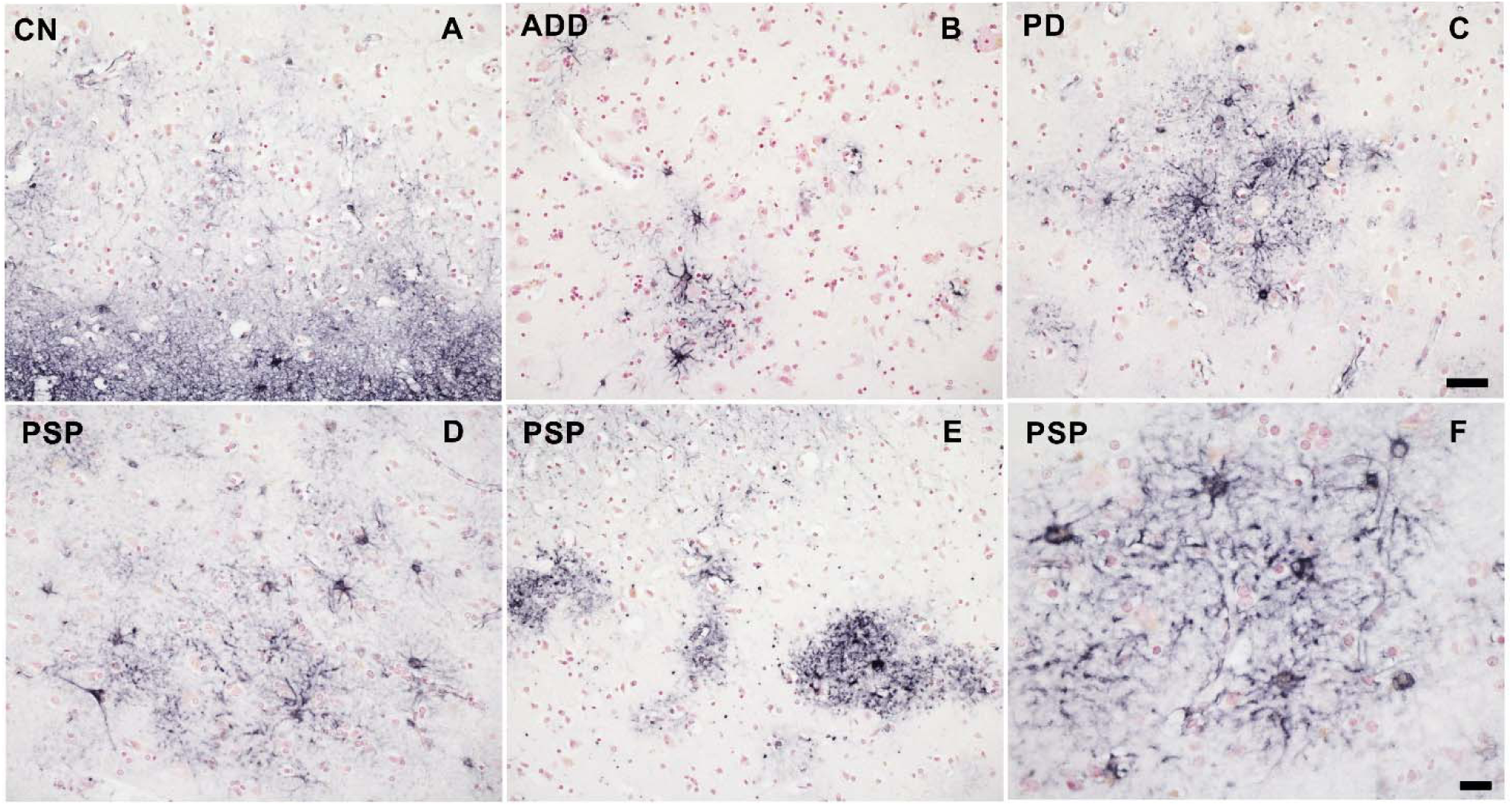
Frontal cortex tissue adjacent to the sample collected for whole-cell-dissociated-suspensions stained for GFAP showed an increased number of reactive astrocytes in PSP cases, especially when compared to controls, ADD and PD.

**Table 1:** Demographics and pathological summary. # p < 0.05 for group comparisons; * p < 0.05 for post-test comparison. n: number of cases; SD: standard deviations; PMI, postmortem interval in hours; CN: cognitively unimpaired low pathology controls; ILBD: incidental Lewy body; PSP: progressive supranuclear palsy; PD: Parkinson’s disease; ADD: Alzheimer’s disease dementia; RIN: RNA Integrity Number

| Diagnosis (n) | Age (SD) | PMI (SD) | LB Total (SD)<sup>#</sup> | Tau Total (SD)<sup>#</sup> | Plaque Total (SD)<sup>#</sup> | PSP & Tau pathology (SD)<sup>#</sup> | RIN (SD) |
| --- | --- | --- | --- | --- | --- | --- | --- |
| CN (3) | 89 (10) | 2.9 (0.3) | 0 | 6 (.6) | 7 (6) | 6 (0.6) | 9.1 (0.2) |
| ILBD (3) | 86 (9) | 3.1 (0.7) | 13 (2) | 5 (3) | 0.6(1) | 5 (3) | 9.5 (0.1) |
| PSP (5) | 89 (7) | 4.4 (1.1) | 2 (4) | 9 (1) | 7 (6) | 27 (9)* | 8.6 (0.7) |
| PD (6) | 82 (14) | 3.7 (0.6) | 31 (5)* | 5 (1) | 0.5 (1) | 5 (1) | 9.2 (0.4) |
| ADD (4) | 87 (4) | 3.2 (1.1) | 0 | 10 (3)* | 12 (3)* | 10 (3) | 9.0 (0.3) |

**Table 2:** Differential gene expression found from each comparison. CN: cognitively unimpaired low pathology controls; ILBD: incidental Lewy body; PSP: progressive supranuclear palsy; PD: Parkinson’s disease; ADD: Alzheimer’s disease dementia.

| Comparison (number of samples) | No. of DEGs |
| --- | --- |
| PSP (5) vs. CN(3) | 1,968 (1,933 upregulated, 35 downregulated) |
| PSP (5) vs. ADD (4) | 89 (88 upregulated, 1 downregulated) |
| PSP (5) vs. PD (6) | 243(4 upregulated, 239 downregulated) |
| ADD (4) vs. CN (3) | 754 (749 upregulated, 5 downregulated) |
| ADD (4) vs. PD (6) | 4 (1 upregulated 3 downregulated) |
| PD (6) vs. CN (3) | 1,150 (53 upregulated, 1,097 downregulated) |
| PD (6) vs. ILBD (3) | 150 (135 upregulated, 15 downregulated) |
| ILBD (3) vs. CN (3) | 1 (downregulated) |
| PSP+PD+ADD (18) vs. CN +ILBD (6) | 503(364 upregulated, 139 downregulated) |

### PSP comparisons

When PSP genes were compared to CN, we found a total of 1,933 upregulated genes and 35 downregulated genes, while expression of UBC, GFAP, MTURN, GLUL, EEF1A1, MAP1A and MAP1B were some of the most significant upregulated *g*enes with over 8 log2(FC) > N changes (Figure 3A). When PSP was compared to ADD we observed a total of 89 genes dysregulated, with 88 of those being upregulated. Some of the most upregulated genes in PSP when compared to ADD were GFAP, CDR1, BCYRN1, and DUSP26 (Figure 3B). When PSP was compared to PD, there were a total of 243 genes dysregulated, with 239 of those being downregulated in PSP, particularly NAT8L, FTO and TUBB2A (Figure 3C).

**Figure 3.**
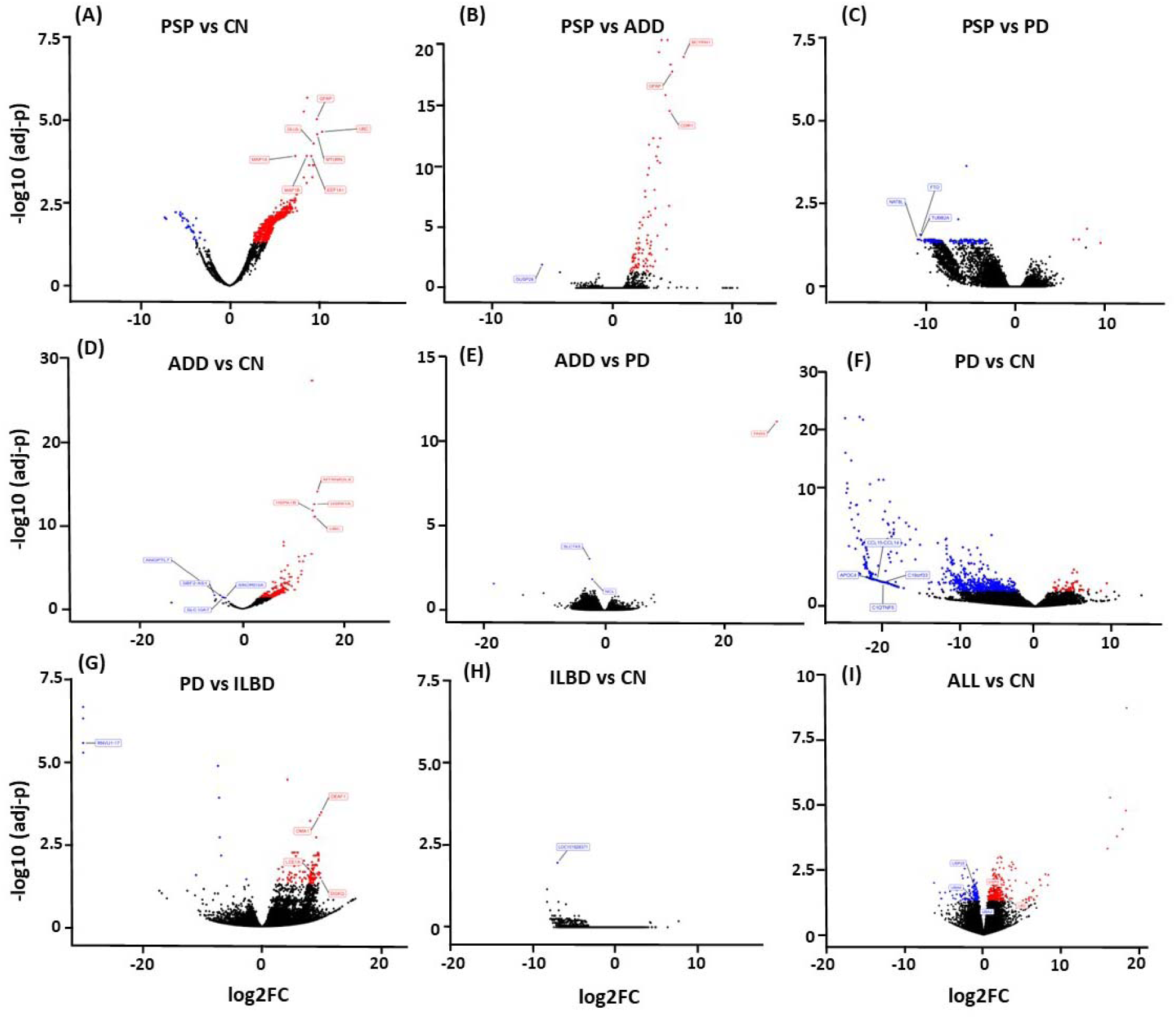
Volcano Plots show dysregulated genes obtained from differential expression analysis comparing gene expression from each diseased group to CN, as well as disease comparison. A) PSP cases had the largest number of dysregulated genes when compared to CN with a total of 1,933 upregulated genes (red) and 35 downregulated genes (blue). B) When PSP was compared to ADD we observed a total of 89 genes dysregulated, C) and when compared to PD 243 genes. D) ADD comparison to CN resulted in 754 genes dysregulated, E) and only 4 genes when compared to PD. F) While PD comparison to CN resulted in,1,150 dysregulated genes, G)135 when PD was compared to ILBD. H) ILBD only had one gene dysregulated when compared to CN, and I) all disease cases showed a total of 501 genes when all cases were combined and compared to CN.

Pathway enrichment analysis revealed the differential processes associated with astrocytes. DEGs obtained from the PSP and CN cases comparison showed an enrichment of dysregulated genes associated with multiple biological pathways, some of the most important involving synaptic pathways, chemokine signaling, protein modification processes and proteolysis (Figure 4A), while the DEGs found from the PSP and ADD cases comparison suggest dysregulation of cellular senescence, cytokine-cytokine receptor, Inositol phosphate metabolism and other cellular processes and interactions (Figure 4B). The PD vs PSP comparison highlights dysregulated genes associated with axonogenesis, glial cell differentiation, histone modification, mitochondrion organization, neurogenesis, pyrophosphatase activity, and regulation of plasma membrane bounded cell projection organization (Complete list in Supplement Table 2).

**Figure 4.**
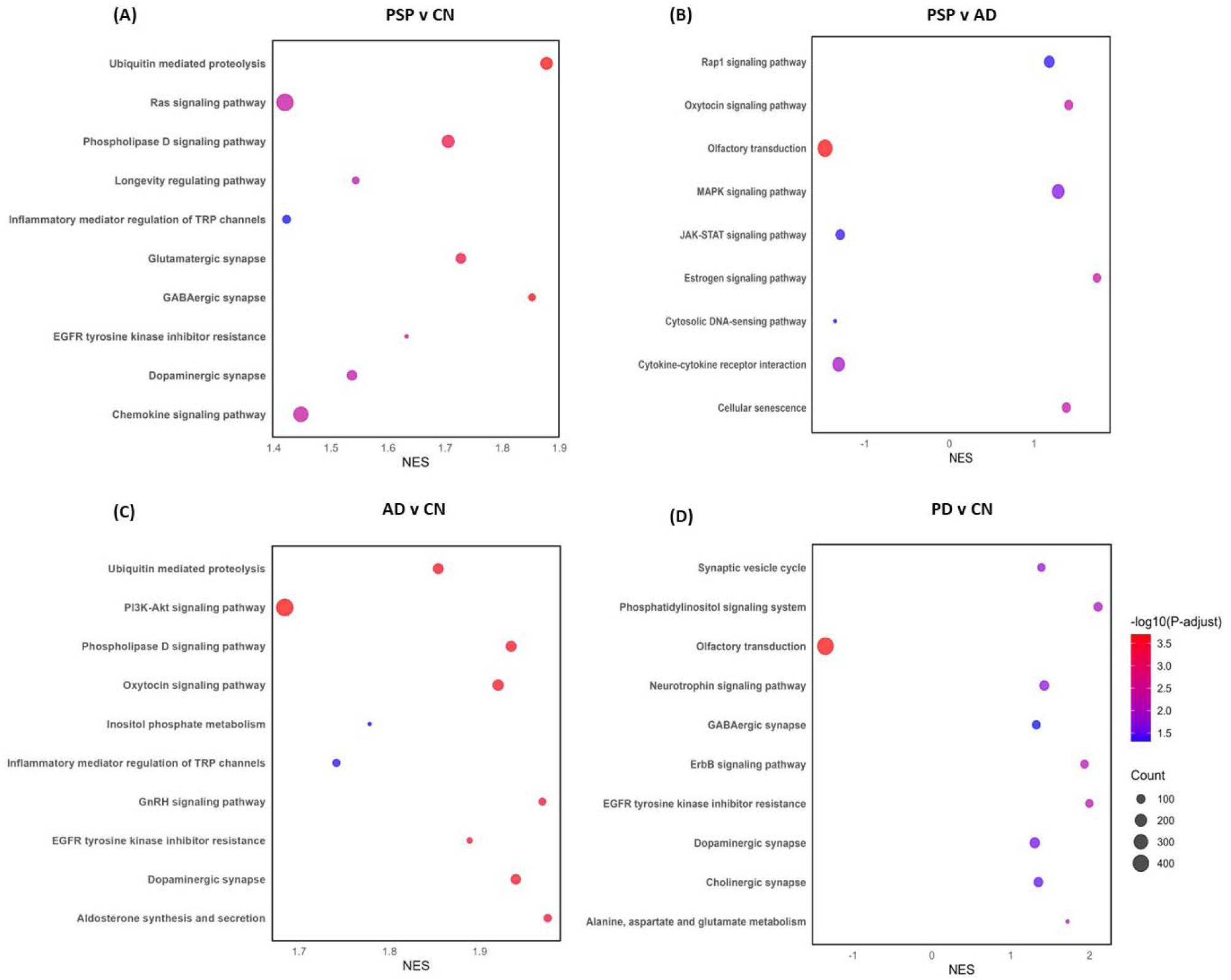
Pathway enrichment analysis obtained from differential expression analysis comparing gene expression from each diseased group to CN, as well as disease comparison. A) PSP and CN cases comparison showed an enrichment of dysregulated genes associated with multiple biological pathways, some of the most important involving synaptic pathways, chemokine signaling, protein modification processes and proteolysis, while when compared to AD we also observed dysregulation of cellular senescence, cytokine-cytokine receptor, Inositol phosphate metabolism and other cellular processes and interactions B). C) ADD shows dysregulation of synapses, proteolysis, and signaling pathways when compared to CN. D) While PD compared to CN showed dysregulation of translocation of olfactory receptors, synapses, synaptic vesicle cycle and neurotrophic signaling pathways.

### ADD comparison

When ADD was compared to CN we found a total of 754 genes dysregulated, 749 of those were upregulated genes and 5 downregulated (SBF2-AS1, LOC100506675, ANGPTL7, SLC10A7, SNORD3A). UBC, MTRNR2L8, HSPA1A, and HSPA1B were the most significantly changed genes in ADD with comparison to controls. Interestingly, comparing PD with ADD showed dysregulation of only 4 genes, PAR4, SLC7A5, MIR548AN and NCL. Thrombin receptor PAR4 was the only gene that was upregulated in ADD when compared to PD, however the dysregulation was one of the largest observed in our comparisons, 28 log2(FC) > N. Pathway enrichment analysis of the DEGs in ADD highlighted dysregulation of synapses, proteolysis, and signaling pathways (Figure 4C).

### PD comparison

When comparing PD astrocytes to CN we found a total of 1,150 dysregulated genes with 53 genes showing upregulation (UBC, HSFY1, OR10J3 and EEF1A2) and 1,097 downregulation. Multiple microRNAs were highly down regulated, showing over 20 log2(FC) > N changes. Other genes highly downregulated included multiple chemokines, multiple immunoglobins, DRD4 and GDF1. Pathway enrichment analysis of DEGs showed dysregulation of translocation of olfactory receptors, synapses, synaptic vesicle cycle and neurotrophic signaling pathways (Figure 4D). Another important comparison made was PD to ILBD. This comparison showed that a total of 135 genes were upregulated and 15 downregulated in PD when compared to possible prodromal cases (ILBD). Some of the most affected genes were DEAF1, DGKQ, OMA1 and ATG101, all showing an upregulation of 9 log2(FC) > N. Enriched pathways were very similar to those observed in PD vs CN, however, both comparisons only have 5 dysregulated genes in common, RPL18A, PPDPF, LCE1A, SNORD38B and RNVU1-17.

### All diseased cases vs CN

To better understand if there are astrocyte genes commonly dysregulated in all neurodegenerative disorder used in this study, we compared the list of dysregulated genes for each disease vs CN comparison. We found that PSP vs CN and ADD vs CN share 669 common dysregulated genes while PD-CN vs ADD-CN share the least number of dysregulated genes with only 10 genes in common (UBC, SBF2-AS1, RN7SL1, RN7SL2). Every gene commonly dysregulated (n=691; comparison #1) was further analyzed for protein–protein interactions (PPI) network using the STRING database. We found 541 nodes and 4952 edges, the overlapping analysis identified five common hub genes (ACTB, EGFR, CALM3, HSP90AB1, UBC; Figure 5).

**Figure 5.**
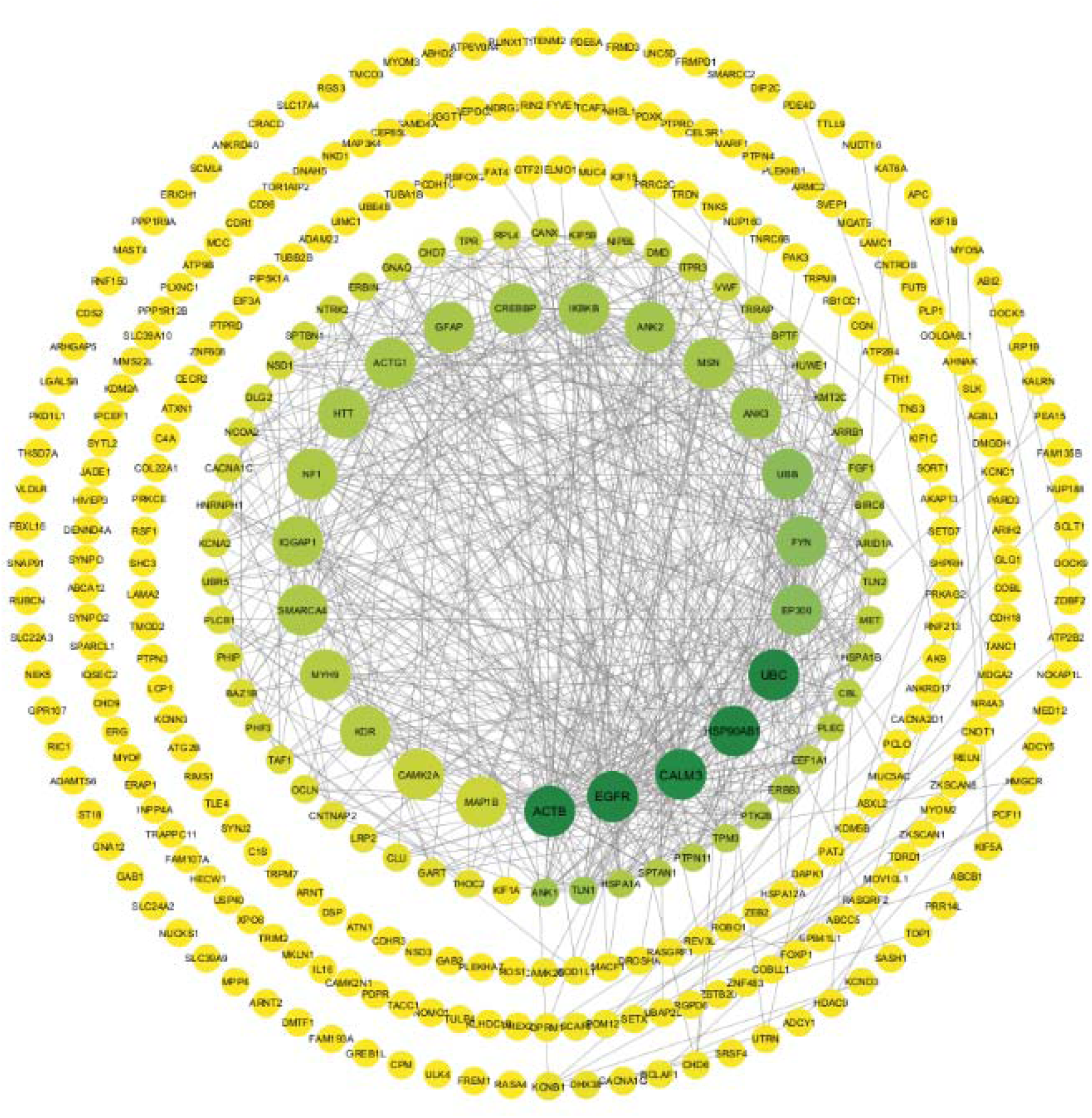
Protein–protein interactions network from a list of 691 common dysregulated genes for each disease vs CN comparison shows 541 nodes and 4952 edges, the overlapping analysis identified five common hub genes (*dark green*; ACTB, EGFR, CALM3, HSP90AB1, UBC).

Another approach for looking at common genes was to combine all disease cases and compare them all with the non-symptomatic cases (CN and ILBD together) using another differential expression analysis. This comparison resulted in a total of 503 DEGs, 364 upregulated and 139 downregulated (Comparison #2). This suggested again common dysregulation of ubiquitin related genes, with UBC, UBA2, UAB6, USP9Y, USP25, and USP3 in all diseases. This PPI network includes 268 nodes and 1124 edges, while the overlapping analysis identified five common hub genes (UBC, YBX1, UTY, RPL4, HSPA1B; Figure 6). Besides the common astrocytic dysregulation of ubiquitin and related genes, we also identified commonly affected biological pathways, such as synaptic dysregulation, transcriptional regulation, VEGFA-VEGFR2 pathway, and chaperone mediated autophagy just to mention some of the pathways with genes in common.

**Figure 6.**
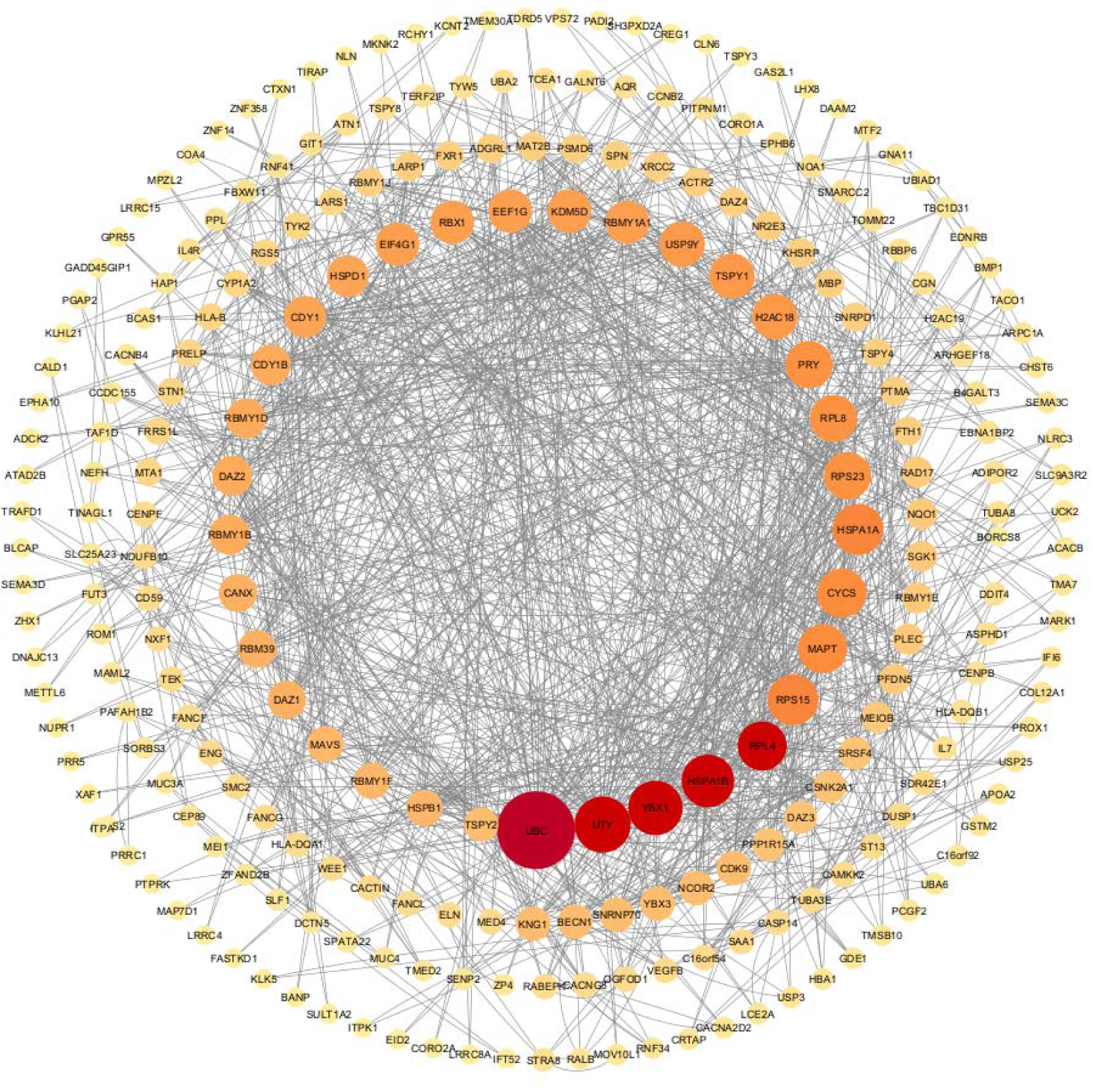
Protein– protein interactions network from a list of 501 common genes from a differential expression analysis combining all disease cases and comparing them all non-symptomatic cases, shows 268 nodes and 1124 edges, while the overlapping analysis identified five commo hub genes (*red;* UBC, YBX1, UTY, RPL4, HSPA1B).

Furthermore, we combined the PPIs from our comparison #1 and #2 using a merge tool of Cytoscape software to find the interconnected and intersected core dysregulated genes and validated the importance of predicted hub genes from each list. UBC, RPL4, and HSPA1A were identified as common hub genes (Figure 7), indicating their central role in the intersected network of dysregulated astrocytic genes across multiple neurodegenerative diseases.

**Figure 7.**
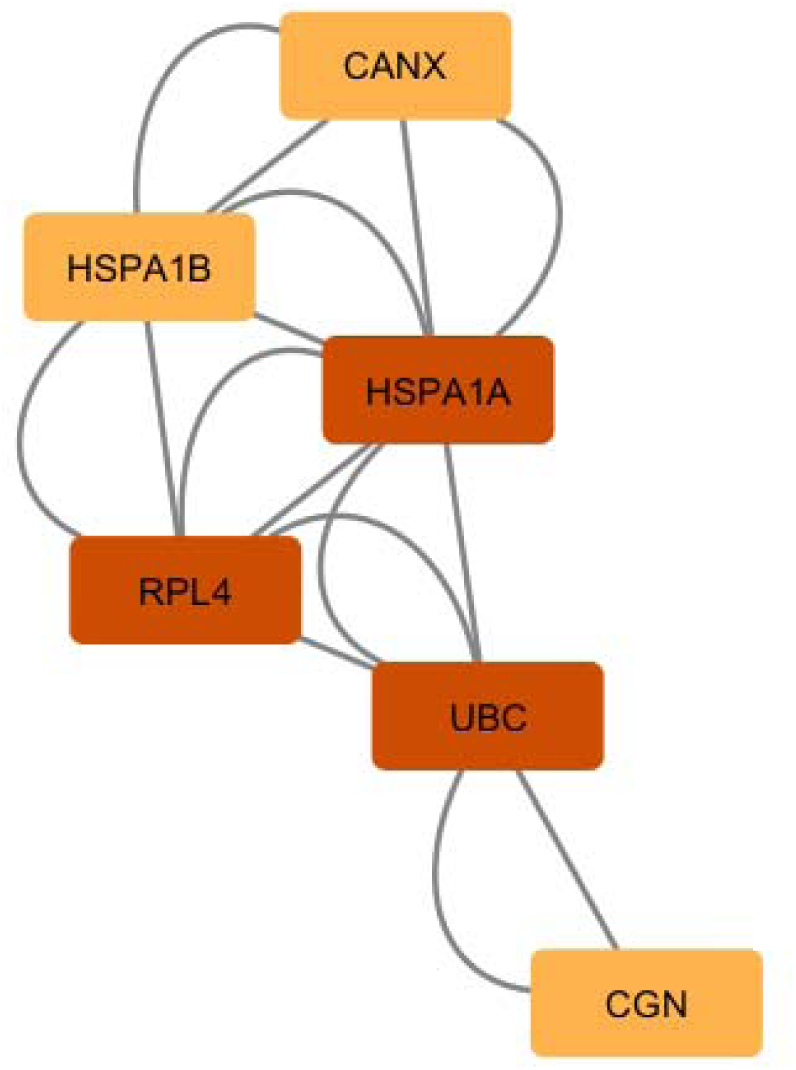
List of common hub genes affected in astrocytes from multiple neurodegenerative diseases. We use the network and hub genes identified in figures 8 and 9 and identified and validated UBC, RPL4, and HSPA1A as important hub genes which networks seems to be commonly affected in PSP, ADD, and PD astrocytes.

The complete dataset for the study is securely posted at Synapse (Project ID: syn64618348).

## DISCUSSION

Multiple studies seem to identify several possible mechanisms of dysfunction in astrocytes from diseased individuals and some have suggested that one therapeutic strategy would be to replace dysfunctional astrocytes with transplanted healthy astrocytes [8, 35]. This was directly tested by Izrael et al (2018) using an amyotrophic lateral sclerosis (ALS) model and indirectly tested by Lee et al (2018) using a PD model (8-9). Izrael et al showed that astrocytes from human embryonic stem cells (hESCs) were safely injected into ALS-mice and rat models and seemed to delay disease onset, slow down disease progression and extend life expectancy. Lee et all injected astrocytes as well as neural progenitor cells (NPC) into the midbrain of a PD-rat model and showed that TH+ cells in the striatum were more numerous, and exhibited healthier and more mature neuronal maturity than those in the control grafts; treated rats showed dramatic behavioral restoration compared to those just treated with NPC injections or shams[8, 36]. Our results support this rationale, as we showed how astrocytes of common neurodegenerative disorders seems to be universally dysregulated when compared to aging normal individuals, suggesting that targeting astrocyte dysfunction could be beneficial for common neurodegenerative diseases.

Our main goal was to study human astrocyte-enriched whole cell suspensions. We are using a unique approach of whole cell analysis rather than individual nuclei analysis for a couple of reasons. We hypothesize that we will be able to analyze a larger number of transcripts by using whole cell compared to nuclei isolation [11, 37–40]. In addition, we believe that by grouping or enriching a population with only one cell type, we should be able to capture smaller changes that might be only present in that type of cell. Whole-homogenate analysis can give completely misleading results, as any biochemical constituent that is selectively localized to the depleted cells will appear to be “down-regulated”, whereas in fact it has most likely been lost only as an “innocent bystander”. Also, a relevant loss or increase might be completely missed, if the biochemical entity is found in many cell types, diluting the ‘lost” signal from the cell of interest, especially if that cell type is uncommon or rare.

We analyzed RNA transcript changes in PSP, ADD and PD human astrocytes when compared to controls. All diseased group comparisons showed prominent dysregulation of genes that are involved in synaptic pathways, metabolic regulation, gene regulation, dephosphorylation and protein modification processes. Perhaps not surprisingly, given the glial tauopathy present in PSP, the most dysregulated astrocyte-enriched populations were those derived from PSP cases. It is well known that astrocytes of PSP cases have aggregated 4-repeat tau that seems to increase their reactivity. In this study we observed that when compared to controls 1,933 upregulated genes and 35 downregulated. Some of the most upregulated genes were astrocyte marker genes such as GFAP and GLUL [41, 42], but also genes associated with protein phosphorylation regulation such as microtubule-associated proteins MAP1A and MAP1B, but not MAPT, which was previously reported to be downregulated in PSP astrocytes [3, 43–46]. Most of the downregulated genes in this comparison were mostly micro RNAs or genes not yet studied. Remarkably, GFAP upregulation was also observed in PSP when the group was compared to CN or ADD but not when compared to PD was compared to any of the groups. This finding was interesting as one would have expected that ADD astrocytes would be more upregulated than PD astrocytes [6, 11, 42]. However, one of our study limitations is the small number of cases and high comorbidity in each group that perhaps prevented us from reaching significance in some of these comparisons.

Other dysregulated genes that are worth mentioning are genes that could explain possible astrocytic dysfunction in ADD, for example, downregulation of SLC10A7 and SNORD3A, which respectively are genes associated with intracellular calcium regulation, and reduced resistance to oxidative and ER stress, as well as upregulations of heat shock HSPA1A, and HSPA1B, proteins associated with apoptosis and ubiquitin-proteasome pathway [39, 47–49]. When comparing PD with control, there were down regulations of many microRNA as well as of many chemokines and immunoglobins genes, suggesting a suppression of inflammatory response in this group which has been previously suggested by others. In this study we also included ILBD cases, a group of subjects that never had parkinsonism during life, but for whom autopsy confirmed the presence of brain a-synuclein pathology and hence represent a probable prodromal stage of PD. Changes observed in this group might suggest astrocytic dysfunction at early stages of a-synuclein pathology. The comparison of ILBD to control only showed the downregulation of one uncharacterized RNA gene, but when the groups were compared to PD we were able to observe up regulation of OMA1, a mitochondrial enzyme that plays a key role in regulating mitochondrial morphology and stress signaling, as well as autophagy-related protein 101 (ATG101) suggesting an early compensatory attempt [2, 50–53]. Another upregulated gene that caught our attention was DGKQ, a diacylglycerol kinase (DGK) that has been proposed as a PD risk gene. DGKs seems to be associated to diverse biological events such as growth factor/cytokine-dependent cell proliferation and motility, immune responses, and glucose metabolism; multiple recent studies suggest that it could be a good target for therapeutic intervention [54–56].

STRING was used to identify the portrait genes with the highest level of interaction with one another as this could provide insight into synergistic effects of altered expression. This analysis emphasized three main machineries involved in protein degradation that seem to be affected in all neurodegenerative enriched astrocytes, namely autophagy, calcium signal transduction pathway and the ubiquitin proteasome system (UPS). Autophagy involves lysosomal degradation, and a large component is regulated by chaperone proteins, while Ca2+ is a universal and versatile signaling molecule that controls many cellular processes, including neurotransmission, cell metabolism, cell death and organelle communication, including endoplasmic reticulum, mitochondria, Golgi complex, and lysosomes [11, 57]. UPS consists of two key components: the ubiquitination system, which selects and targets proteins towards degradation by ubiquitinating them, and the proteasome. It is well known that ubiquitination-mediated control contributes to the maintenance of cellular homeostasis, and dysregulation of ubiquitination reactions plays a relevant role in the pathogenic states of neurodegenerative diseases [58–60]. Previous research work described how important this system is in glia cells, but none have clearly identified how dysregulated this system is in astrocytes derived from common neurodegenerative diseases [61]. Ubiquitination can alter the molecular functions of tagged substrates with respect to protein turnover, biological activity, subcellular localization, or protein–protein interaction. The UPS has been implicated in pathways that regulate neurotransmitter release, synaptic membrane receptor turnover and synaptic plasticity and as a result, affects a wide variety of cellular processes, with chronic overexpression potentially inducing synaptic dysfunction in neurons. Our results strongly support the rationale that modulation of astrocytes should be further evaluated as a potential therapeutic strategy [62, 63].

## Data Availability

all data will be posted in Synapse (Project ID syn64618348).

https://help.nf.synapse.org/NFdocs/about-data-sharing

## Notes

### Competing Interest Statement

The authors have declared no competing interest.

### Funding Statement

The Arizona Study of Aging and Neurodegenerative Disorders Brain and Body Donation Program at Banner Sun Health Research Institute, Sun City, Arizona has been supported by the National Institute of Neurological Disorders and Stroke (U24 NS072026 National Brain and Tissue Resource for Parkinson Disease and Related Disorders), the National Institute on Aging (P30 AG019610 and P30AG072980, Arizona Alzheimer Disease Center), the Arizona Department of Health Services (contract 211002, Arizona Alzheimer Research Center), the Arizona Biomedical Research Commission (contracts 4001, 0011, 05-901 and 1001 to the Arizona Parkinson Disease Consortium) and the Michael J. Fox Foundation for Parkinson Research.

### Author Declarations

The study was conducted in accordance with the Declaration of Helsinki and approved by the Institutional Review Board of WCG for studies involving humans. Study Number 1132516.

### Summary of Updates

Removal of duplicated figures. Upload mistake.

## REFERENCES

1. Booth, H.D.E., W.D. Hirst, and R. Wade-Martins, The Role of Astrocyte Dysfunction in Parkinson’s Disease Pathogenesis. Trends Neurosci, 2017. 40(6): p. 358–370.

2. Sorrentino, Z.A., B.I. Giasson, and P. Chakrabarty, *alpha-Synuclein and astrocytes: tracing the pathways from homeostasis to neurodegeneration in Lewy body disease*. Acta Neuropathol, 2019. 138(1): p. 1–21.

3. Fan, L.Y., et al., Single-nucleus transcriptional profiling uncovers the reprogrammed metabolism of astrocytes in Alzheimer’s disease. Front Mol Neurosci, 2023. 16: p. 1136398.

4. Miyazaki, I. and M. Asanuma, Neuron-Astrocyte Interactions in Parkinson’s Disease. Cells, 2020. 9(12).

5. Benarroch, E.E., Astrocyte signaling and synaptic homeostasis: I: Membrane channels, transporters, and receptors in astrocytes. Neurology, 2016. 87(3): p. 324–30.

6. Perez-Nievas, B.G. and A. Serrano-Pozo, Deciphering the Astrocyte Reaction in Alzheimer’s Disease. Front Aging Neurosci, 2018. 10: p. 114.

7. Song, Y.J., et al., Degeneration in different parkinsonian syndromes relates to astrocyte type and astrocyte protein expression. J Neuropathol Exp Neurol, 2009. 68(10): p. 1073–83.

8. Song, J.J., et al., Cografting astrocytes improves cell therapeutic outcomes in a Parkinson’s disease model. J Clin Invest, 2018. 128(1): p. 463–482.

9. Wang, C., et al., Astrocyte dysfunction in Parkinson’s disease: from the perspectives of transmitted alpha-synuclein and genetic modulation. Transl Neurodegener, 2021. 10(1): p. 39.

10. Boisvert, M.M., et al., The Aging Astrocyte Transcriptome from Multiple Regions of the Mouse Brain. Cell Rep, 2018. 22(1): p. 269–285.

11. Wang, X., et al., Alzheimer’s disease and progressive supranuclear palsy share similar transcriptomic changes in distinct brain regions. J Clin Invest, 2022. 132(2).

12. Chai, H., et al., Neural Circuit-Specialized Astrocytes: Transcriptomic, Proteomic, Morphological, and Functional Evidence. Neuron, 2017. 95(3): p. 531–549 e9.

13. Tagliafierro, L., et al., Gene Expression Analysis of Neurons and Astrocytes Isolated by Laser Capture Microdissection from Frozen Human Brain Tissues. Front Mol Neurosci, 2016. 9: p. 72.

14. Beach, T.G., et al., Arizona Study of Aging and Neurodegenerative Disorders and Brain and Body Donation Program. Neuropathology, 2015. 35(4): p. 354–389.

15. Beach, T.G., et al., The Sun Health Research Institute Brain Donation Program: description and experience, 1987-2007. Cell Tissue Bank, 2008. 9(3): p. 229–245.

16. Beach, T.G., et al., Arizona Study of Aging and Neurodegenerative Disorders and Brain and Body Donation Program. Neuropathology, 2015. 34(4): p. 354–389.

17. Braak, H. and E. Braak, Neuropathological stageing of Alzheimer-related changes. Acta Neuropathol, 1991. 82(4): p. 239–259.

18. Braak, H. and E. Braak, Demonstration of amyloid deposits and neurofibrillary changes in whole brain sections. Brain Pathol, 1991. 1(3): p. 213–6.

19. Aging, N.I.o., Consensus recommendations for the postmortem diagnosis of Alzheimer’s disease. The National Institute on Aging, and Reagan Institute Working Group on Diagnostic Criteria for the Neuropathological Assessment of Alzheimer’s Disease. Neurobiol. Aging, 1997. 18(4 Suppl): p. S1–S2.

20. Hyman, B.T. and J.Q. Trojanowski, Consensus recommendations for the postmortem diagnosis of Alzheimer disease from the National Institute on Aging and the Reagan Institute Working Group on diagnostic criteria for the neuropathological assessment of Alzheimer disease. J. Neuropathol. Exp. Neurol, 1997. 56(10): p. 1095–1097.

21. Mirra, S.S., The CERAD neuropathology protocol and consensus recommendations for the postmortem diagnosis of Alzheimer’s disease: a commentary. Neurobiol. Aging, 1997. 18(4 Suppl): p. S91–S94.

22. Montine, T.J., et al., National Institute on Aging-Alzheimer’s Association guidelines for the neuropathologic assessment of Alzheimer’s disease: a practical approach. Acta Neuropathol, 2012. 123(1): p. 1–11.

23. Beach, T.G., et al., Unified staging system for Lewy body disorders: correlation with nigrostriatal degeneration, cognitive impairment and motor dysfunction. Acta Neuropathol, 2009. 117(6): p. 613–634.

24. Kovacs, G.G., et al., Distribution patterns of tau pathology in progressive supranuclear palsy. Acta Neuropathol, 2020. 140(2): p. 99–119.

25. Serrano, G.E., et al., Whole-Cell Dissociated Suspension Analysis in Human Brain Neurodegenerative Disease: A Pilot Study. medRxiv, 2021: p. 2021.01.08.21249455.

26. Dobin, A., et al., STAR: ultrafast universal RNA-seq aligner. Bioinformatics, 2013. 29(1): p. 15–21.

27. Chandramohan, R., et al. Benchmarking RNA-Seq quantification tools. in 2013 35th Annual International Conference of the IEEE Engineering in Medicine and Biology Society (EMBC). 2013. IEEE.

28. Ewels, P., et al., MultiQC: summarize analysis results for multiple tools and samples in a single report. Bioinformatics, 2016. 32(19): p. 3047–3048.

29. Love, M.I., W. Huber, and S. Anders, Moderated estimation of fold change and dispersion for RNA-seq data with DESeq2. Genome biology, 2014. 15: p. 1–21.

30. Mathys, H., et al., Single-cell transcriptomic analysis of Alzheimer’s disease. Nature, 2019. 570(7761): p. 332–337.

31. Piras, I.S., et al., Olfactory bulb and amygdala gene expression changes in subjects dying with COVID-19. medRxiv, 2021.

32. Wu, T., et al., *clusterProfiler 4.0:* A universal enrichment tool for interpreting omics data. The innovation, 2021. 2(3).

33. Bajpai, A.K., et al., Systematic comparison of the protein-protein interaction databases from a user’s perspective. Journal of Biomedical Informatics, 2020. 103: p. 103380.

34. Kohl, M., S. Wiese, and B. Warscheid, Cytoscape: software for visualization and analysis of biological networks. Data mining in proteomics: from standards to applications, 2011: p. 291–303.

35. Gorshkov, K., et al., Astrocytes as targets for drug discovery. Drug Discov Today, 2018. 23(3): p. 673–680.

36. Izrael, M., et al., Safety and efficacy of human embryonic stem cell-derived astrocytes following intrathecal transplantation in SOD1(G93A) and NSG animal models. Stem Cell Res Ther, 2018. 9(1): p. 152.

37. Whitney, K., et al., Single-cell transcriptomic and neuropathologic analysis reveals dysregulation of the integrated stress response in progressive supranuclear palsy. bioRxiv, 2023.

38. Dai, D.L., M. Li, and E.B. Lee, Human Alzheimer’s disease reactive astrocytes exhibit a loss of homeostastic gene expression. Acta Neuropathol Commun, 2023. 11(1): p. 127.

39. Caldi Gomes, L., et al., Multi-omic landscaping of human midbrains identifies disease-relevant molecular targets and pathways in advanced-stage Parkinson’s disease. Clin Transl Med, 2022. 12(1): p. e692.

40. Sadick, J.S., et al., Astrocytes and oligodendrocytes undergo subtype-specific transcriptional changes in Alzheimer’s disease. Neuron, 2022. 110(11): p. 1788–1805 e10.

41. Monterey, M.D., et al., The Many Faces of Astrocytes in Alzheimer’s Disease. Front Neurol, 2021. 12: p. 619626.

42. Kim, J., et al., Pathological phenotypes of astrocytes in Alzheimer’s disease. Exp Mol Med, 2024. 56(1): p. 95–99.

43. Briel, N., et al., Single-nucleus chromatin accessibility profiling highlights distinct astrocyte signatures in progressive supranuclear palsy and corticobasal degeneration. Acta Neuropathol, 2022. 144(4): p. 615–635.

44. Farrell, K., et al., Genetic, transcriptomic, histological, and biochemical analysis of progressive supranuclear palsy implicates glial activation and novel risk genes. Nat Commun, 2024. 15(1): p. 7880.

45. Jackson, R.J., et al., Astrocyte tau deposition in progressive supranuclear palsy is associated with dysregulation of MAPT transcription. Acta Neuropathol Commun, 2024. 12(1): p. 132.

46. Forrest, S.L., et al., Cell-specific MAPT gene expression is preserved in neuronal and glial tau cytopathologies in progressive supranuclear palsy. Acta Neuropathol, 2023. 146(3): p. 395–414.

47. Chen, Y., et al., Identification of novel hub genes for Alzheimer’s disease associated with the hippocampus using WGCNA and differential gene analysis. Front Neurosci, 2024. 18: p. 1359631.

48. Park, H.K., et al., MPTP-induced model of Parkinson’s disease in heat shock protein 70.1 knockout mice. Mol Med Rep, 2012. 5(6): p. 1465–8.

49. Singh, R., S. Kolvraa, and S.I. Rattan, Genetics of human longevity with emphasis on the relevance of HSP70 as candidate genes. Front Biosci, 2007. 12: p. 4504–13.

50. Liu, Y.T., et al., Loss of CHCHD2 and CHCHD10 activates OMA1 peptidase to disrupt mitochondrial cristae phenocopying patient mutations. Hum Mol Genet, 2020. 29(9): p. 1547–1567.

51. Rivera-Mejias, P., et al., The mitochondrial protease OMA1 acts as a metabolic safeguard upon nuclear DNA damage. Cell Rep, 2023. 42(4): p. 112332.

52. Guo, X., et al., Mitochondrial stress is relayed to the cytosol by an OMA1-DELE1-HRI pathway. Nature, 2020. 579(7799): p. 427–432.

53. Malik, B.R., et al., Autophagic and endo-lysosomal dysfunction in neurodegenerative disease. Mol Brain, 2019. 12(1): p. 100.

54. Gu, X.J., et al., Expanding causal genes for Parkinson’s disease via multi-omics analysis. NPJ Parkinsons Dis, 2023. 9(1): p. 146.

55. Zhou, S., et al., Brain Proteome-Wide and Transcriptome-Wide Asso-ciation Studies, Bayesian Colocalization, and Mendelian Randomization Analyses Reveal Causal Genes of Parkinson’s Disease. J Gerontol A Biol Sci Med Sci, 2023. 78(4): p. 563–568.

56. Sakane, F., S. Mizuno, and S. Komenoi, Diacylglycerol Kinases as Emerging Potential Drug Targets for a Variety of Diseases: An Update. Front Cell Dev Biol, 2016. 4: p. 82.

57. Hill, M.A. and S.C. Gammie, Alzheimer’s disease large-scale gene expression portrait identifies exercise as the top theoretical treatment. Sci Rep, 2022. 12(1): p. 17189.

58. Zhang, H., et al., USP3: Key deubiquitylation enzyme in human diseases. Cancer Sci, 2024. 115(7): p. 2094–2106.

59. Zhang, Y., et al., Bioinformatic Analysis and Experimental Validation of Ubiquitin-Proteasomal System-Related Hub Genes as Novel Biomarkers for Alzheimer’s Disease. J Integr Neurosci, 2023. 22(6): p. 138.

60. Wu, X., et al., Elucidating Microglial Heterogeneity and Functions in Alzheimer’s Disease Using Single-cell Analysis and Convolutional Neural Network Disease Model Construction. Sci Rep, 2024. 14(1): p. 17271.

61. Jansen, A.H., E.A. Reits, and E.M. Hol, The ubiquitin proteasome system in glia and its role in neurodegenerative diseases. Front Mol Neurosci, 2014. 7: p. 73.

62. Spano, D. and G. Catara, Targeting the Ubiquitin-Proteasome System and Recent Advances in Cancer Therapy. Cells, 2023. 13(1).

63. Huang, X. and V.M. Dixit, Drugging the undruggables: exploring the ubiquitin system for drug development. Cell Res, 2016. 26(4): p. 484–98.

